# A SIX WEEK PERFORMANCE BASED CONCENTRIC ONLY RESISTANCE TRAINING PROTOCOL INCREASES STRENGTH IN OLDER ADULTS

**DOI:** 10.1101/2025.06.19.25329807

**Authors:** Amanda L. Rotondo, Eric Lam, Gordon Lam, Rebecca Macdonald, Ka-Wah Ng, Maryann Coughlin, Fiona Ho

## Abstract

**Background and Purpose:** Resistance training has been widely studied in the older adult population as it relates to strength and function, however, there is limited research on the use of concentric resistance-only training using hydraulic exercise equipment. This study aimed to determine the effects of a 6 week performance-based concentric only resistance training protocol on isometric muscle strength, balance, fall risk, and perceived lower extremity function in older adults.

**Methods:** Thirteen older adults (74.1±7.1 years, 6 male, 7 female) participated in this study. The intervention consisted of a concentric resistance-only circuit training program using hydraulic exercise machines equipped with Gym Tonic software twice per week for 6 weeks. Movements included knee flexion/extension, trunk flexion/extension, hip abduction/adduction, chest press/row, and leg press/squat. The software automatically adjusted the resistance based on user performance. Outcome measures included isometric muscle strength, Berg Balance Scale (BBS), Timed Up and Go (TUG), and Lower Extremity Functional Scale (LEFS). Isometric muscle strength data were analyzed using paired t-tests and functional outcome measures were analyzed using Wilcoxon signed-rank test. Bonferroni correction was used to set a new alpha level of 0.0038.

**Results:** Significant increases in isometric muscle strength were observed for knee flexion (p=0.002), knee extension (p=0.001), chest press (p=0.001), and leg press (p<0.001). No statistically significant differences (p>0.0038) or clinically (Minimal Detectable Change (MDC), Minimal Clinically Important Difference (MCID)) significant differences were observed for BBS, TUG, and LEFS.

**Discussion:** A 6-week intervention of concentric resistance only training using hydraulic resistance training equipment significantly improved isometric muscle strength of knee flexion, knee extension, chest press, and leg press in older adults. While no significant differences were found for BBS, LEFS, and TUG scores, potentially due to the ceiling effect of participant baseline scores and short duration of intervention, positive trends were noted.

**Conclusion:** Physical Therapists should consider using concentric only resistance training on hydraulic equipment to safely improve motor performance in older adults.

## INTRODUCTION

Preserving physical function and independence is crucial for older adults, particularly given the growing aging global population. Loss of mobility significantly elevates fall risk and diminishes quality of life.^1^ Therefore, the need to maintain functional independence in this population is imperative. To address this, interventions aimed at enhancing mobility and overall health are urgently needed. Strengthening muscle function is a key component of these interventions, as it directly supports independence, fosters better health, and enhances the overall quality of life.^1^

Aging leads to progressive declines in the musculoskeletal system, characterized by reductions in muscle mass, bone density, and cartilage integrity, collectively increasing the risk of frailty and functional impairments.^2,3^ Sarcopenia, the age-related loss of muscle mass and strength, is driven by multiple factors, including mitochondrial dysfunction, increased oxidative stress, and chronic low-grade inflammation.^4, 5^ These cumulative musculoskeletal changes contribute to diminished mobility, increased fall risk, and a decline in the overall quality of life as individuals age.

Resistance training is an effective strategy to counteract age-related decline. It has been shown to promote muscle hypertrophy in older adults.^6,7^ Specifically, resistance training enhances muscle function and strength in individuals over 50, with significant increases observed in both lower and upper body strength after training protocols lasting from 6 to 52 weeks, and training intensities ranging from 40% to 85% of one-repetition maximum (1RM).^1^ Additionally, resistance training increases muscle mass, motor unit recruitment, and firing rate in individuals over 60 years old. Training at 60–85% of an individual’s maximum voluntary strength promotes muscle hypertrophy, while intensities above 85% improve the rate of force development.^8^ Therefore, structured resistance exercise programs enhance muscle power and motor unit recruitment, leading to increased ability to adjust to perturbations and recover from loss of balance.

In addition to increasing muscle mass and power, resistance training has been shown to significantly improve gait and balance parameters in older adults, which are critical for reducing the fall risk and maintaining independence. Resistance training leads to notable improvements in walking speed, stride length, and overall postural stability in older adults following training interventions lasting 6-32 weeks, with 2–3 sessions per week and 8–15 repetitions per set.^9^

Although traditional resistance training has been shown to improve strength and function in older adults, hydraulic resistance training (HRT) presents a promising alternative. HRT includes concentric only contractions, eliminating eccentric contractions, which are known to be associated with muscle damage.^10,11^ While eccentric contractions use higher force and less perceived effort, concentric contractions have been associated with increased muscle strength and improvements in functional outcome measures.^12,13^ Utilizing only concentric contractions during exercise can also reduce joint stress and muscle strain, making it a potentially safer option for this population.^14^

HRT also adapts to the user’s range of motion and automatically halts when input ceases, further enhancing safety. These characteristics make HRT a low-impact and effective choice for older adults, offering key advantages over traditional resistance training methods, such as free weights and weight machines. After participating in a 12-week supervised HRT program, healthy, untrained older adults experienced significant increases in strength.^15^ These findings demonstrate that the motor output improvements achieved through HRT are in line with those observed with traditional weight training methods.

Despite these prior findings, the impact of hydraulic resistance training on various motor functions in older adults has not been studied extensively. Given the safety benefits of HRT, it has the potential to be a valuable intervention for improving balance and reducing fall risk in older adults. The enhanced muscle strength and stability gained through HRT may contribute to better postural control and coordination which are key factors in fall prevention. Therefore, further research into HRT’s role in improving functional mobility and lowering fall risk is essential.

While prior studies on HRT have largely focused on longer intervention periods, this study aimed to determine whether similar functional benefits can be observed after just 6 weeks of training. This study aimed to evaluate the effects of a 6-week concentric only HRT protocol on balance, perceived lower extremity function, fall risk, and muscle strength in older adults. The training program included exercises targeting trunk flexion/extension, knee flexion/extension, chest press, back row, hip abduction/adduction, leg press, and hip flexion. The outcomes assessed included isometric muscle strength, Berg Balance Scale (BBS), Lower Extremity Functional Scale (LEFS), and Timed Up and Go (TUG) test.

We hypothesized that a 6-week concentric only resistance training protocol using HRT may lead to improvements in balance, fall risk, perceived lower extremity function, and muscle strength in older adults, with changes potentially approaching or exceeding the minimal detectable change (MDC) for the BBS, TUG, and LEFS. The results of this study can aid the development of safe concentric resistance only protocols using HRT in older adults and add to the existing body of evidence.

## METHODS

### Ethics Approval

This study was approved by the City University of New York Institutional Review Board (2024-0367-CSI). This study is registered at Clinicaltrials.gov with ID: NCT06353438. All of the participants provided written informed consent.

### Study Design

This was a quasi-experimental study using a within-subject design aimed at assessing the impact of use of concentric only resistance training on HRT equipment on strength and function in older adults over a 6-week period.

### Location

This study was conducted at a senior center in Brooklyn, NY. This facility provides services and programs for individuals aged 60 and above.

### Participants

Participants (n=20) were selected through convenience sampling based on their membership to the senior center. Participants were screened according to the inclusion and exclusion criteria outlined in Table 1. Individuals who were scheduled for evaluations on the HRT machines at the senior center were invited to participate in the study.

**Table 1.** Inclusion and Exclusion Criteria. Table 1 presents the inclusion and exclusion criteria for participation.

| Inclusion Criteria | Exclusion Criteria |
| --- | --- |
| Healthy Adults<br>Age 60+ | Age <60<br>Neurological condition<br>Surgery within last 6 months<br>Current injury<br>Inability to follow directions<br>Inability to participate in physical activity for any medical reason |

### Equipment

This study utilized the FREI Medical FACTUM® line of HRT machines.^16^ The machine circuit included five individual pieces of equipment which were manufactured in Germany. *Figure 1* shows images of the equipment. Training on the machines is concentric/concentric working both agonist and antagonist muscle groups/movements on the same piece of equipment (i.e. knee flexion and knee extension) in one flow of movement.^16^

**Figure 1.**
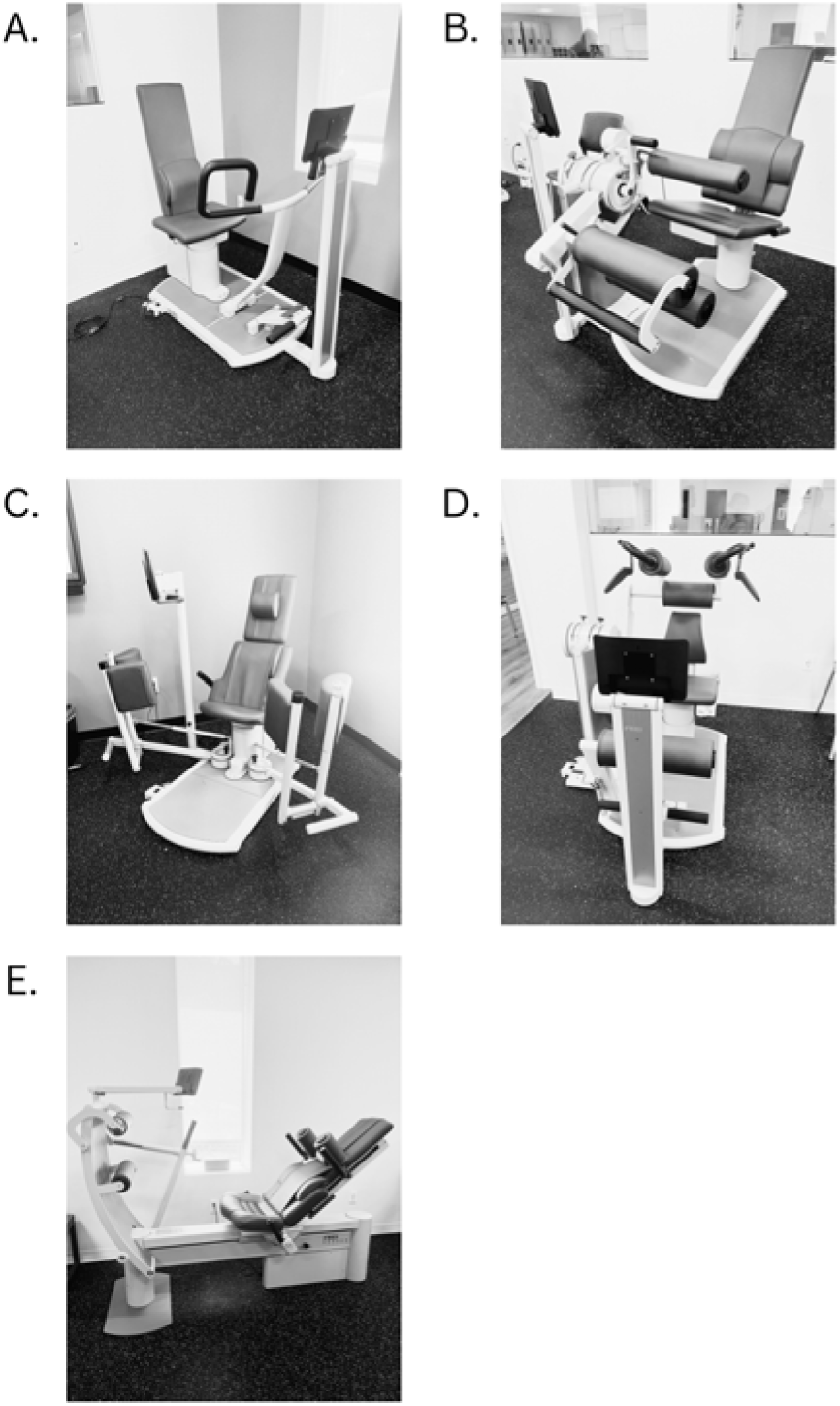
Images of Hydraulic Resistance Training Equipment. A.Chest Press and Row Machine B. Knee Flexion and Knee Extension Machine C. Hip Abduction and Hip Adduction Machine D. Trunk Flexion and Trunk Extension Machine E. Leg Press and Squat Machine

### Software

The HRT machines are equipped with Gym Tonic software, an evidence-based AI software that tests isometric muscle strength and plans, controls, and documents individual sessions.^17,18^ The AI software adapts resistance automatically based on user performance to create progressive overload, and the hydraulic equipment offers smooth stopping at any time, reducing the risk of injury.

### Outcome Measures

The outcome measures utilized during this study were isometric muscle strength (expressed as peak torque) using maximum voluntary contraction testing, the Berg Balance Scale (BBS), Timed Up and Go (TUG), and Lower Extremity Functional Scale (LEFS). Isometric strength testing was performed by a Physical Therapist and Certified Personal Trainer who were trained on the equipment and use of software from their respective companies. The functional outcome measures (BBS, TUG, LEFS) were assessed by a Physical Therapist at initial screening and at the end of the intervention period.

Isometric muscle strength testing was performed on each machine for each motion including trunk flexion and extension, knee flexion and extension, squat and leg press/stand, hip abduction and adduction, and chest press and seated row. Participants were instructed to exert maximal force for 8 seconds against the equipment for each movement, resulting in an isometric contraction. A visual aid for biofeedback on the equipment screen showed the participant how much force they exerted during the contraction. Compensations were corrected and movements were re-tested if needed. Each movement was tested 3 times each and the highest attempt was recorded in Newton-meters (Nm).

The BBS and TUG were chosen for their sensitivity and specificity in identifying fallers and non-fallers (BBS specificity: 75%, TUG sensitivity and specificity: 87%).^19,20^ The LEFS was chosen for its reliability in identifying functional lower extremity limitations (LEFS Test-retest reliability R = 0.94).^21^

Scores on the BBS range from 0-56 points with 14 items scoring from 0-4 points on each item. Scores of 0-20 indicate high fall risk, 21-40 moderate risk, and 41-56 indicates low fall risk.^22^ The minimal detectable change (MDC) for the BBS is 6.5 points (for older adults) and and the minimal clinically important difference (MCID) is 5 points (for patients with early subacute stroke).^23,24^ The TUG is scored based on time; scores of 10 seconds or less indicate normal mobility and scores >13.5 seconds indicate increased risk of falls.^25^ The MDC and MCID for TUG is: 3 seconds (for community dwelling older adults) and 3.4-3.5 seconds (for people with Lumbar Degenerative Disc Disease and people with Parkinson’s Disease, respectively).^26,27,28^ The LEFS is a 20 item self-reported questionnaire with scores ranging from 0-80. Each item on the LEFS ranges from 0-4: 0 (extreme difficulty/unable to perform), 1 (quite a bit of difficulty), 2 (moderate difficulty), 3 (a little bit of difficulty) and 4 (no difficulty).

Scores of 20-40 may indicate moderate to severe functional limitation, 40-61 may indicate moderate functional limitation, and 61-80 may indicate normal or minimal functional limitations.^21^ The MDC and MCID for LEFS are 9 points and 9 points.^29^ MCID values for community dwelling older adults could not be found for the BBS and TUG; therefore reported values reflect those for specific patient populations.

### Study Protocol

At the initial assessment, participants completed the LEFS and were assessed using the BBS and TUG by a Physical Therapist. Following these assessments, participants were issued a personalized access card to use with the HRT equipment and software and were introduced to the circuit of machines. Isometric muscle strength was then measured for each movement.

Following the initial assessment, participants were instructed to utilize the 5 machines in a circuit twice per week for 6 weeks under the supervision of a Physical Therapist and Certified Personal Trainer who were trained on the equipment and use of software from their respective companies. Each participant used the personalized access card on each machine which recorded their workouts, enabling tracking of their participation. During this initial period, attendance was not strictly enforced, and participants were not excluded based on session frequency, whether they attended more or less than the recommended twice per week. After this 6-week period, adherence levels were assessed based on participation data, and participants were excluded if they did not complete at least 6 sessions (approximately 1 session per week) or if they completed more than 13 sessions, to prevent under and over exposure. No limitations were placed on physical activities outside of this study protocol.

The exercise circuit lasted anywhere from 30 to 45 minutes and included concentric movements only against resistance for agonist and antagonist muscles on the same piece of equipment in one flow of movement: trunk flexion and extension, knee flexion and extension, squat and stand/leg press, hip abduction and adduction, and chest press and seated row.

Participants performed 3 sets of 10 repetitions on each machine. The software automatically determined the appropriate resistance in Newtons (N) for the participant based on isometric strength testing and performance on the previous set and in the previous session. The first set was a warmup at 50% of the predetermined resistance level. The second and third sets were set at 100% of the predetermined resistance level. The resistance was automatically adjusted throughout the workout by the software based on individual performance. The software increased the resistance if the participant moved too quickly through the set or decreased the resistance if the user took increased time to complete the set. The software also adjusted the resistance based on the user’s range of motion; if a participant was unable to move through their available range of motion, the resistance was decreased. These automatic adjustments ensure progressive overload while maintaining safety.

Participants were contacted for post-testing 6 weeks after enrollment in the study. They were instructed not to utilize the equipment on the same day as their post-testing session. Post-testing procedures for LEFS, BBS, and TUG were conducted by a Physical Therapist and post-testing for isometric muscle strength was conducted by a Physical Therapist and Certified Personal Trainer trained on the equipment and software.

### Statistical Analysis

The data distribution was assessed for normality using the Shapiro-Wilk test. For comparing 2 paired groups, paired t-tests were used to analyze normally distributed data and Wilcoxon signed-rank test was used to analyze data that was not normally distributed. To account for multiple comparisons (13 tests), the Bonferroni correction was applied, adjusting the significance level to = 0.0038 (0.05/13). Statistical analyses were performed using IBM SPSS Statistics for Windows, version 27 (IBM Corp., Armonk, N.Y., USA).

## RESULTS

### Participants

Twenty individuals were initially enrolled in the study. Three participants withdrew voluntarily, 3 were excluded for not meeting visit requirements, and 1 was removed due to a neurological comorbidity. Therefore, data from 13 participants (6 male, 7 female) with an average age of 74.1 years (SD= 7.1) were included in the final analysis. As shown in Table 2, adherence to the training program was mixed. Only 3 participants followed the prescribed schedule of 2 sessions per week for 6 weeks. Nine participants attended fewer than 12 sessions, whereas 1 participant attended more than 12 sessions.

**Table 2.** Number of Sessions Attended. Table 2 presents number of sessions attended over the 6-week intervention period.

| Subject Study Code | Sessions Attended |
| --- | --- |
| 1 | 12 |
| 2 | 12 |
| 3 | 12 |
| 4 | 7 |
| 5 | 11 |
| 6 | 11 |
| 7 | 13 |
| 8 | 8 |
| 9 | 8 |
| 10 | 6 |
| 11 | 10 |
| 12 | 9 |
| 13 | 7 |
| Total: 13 | Mean ± SD: 9.7 ± 2.32 |

*Notes: SD:Standard Deviation*

### Muscle Strength

The Shapiro-Wilk test indicated that all isometric muscle strength data were normally distributed (p>0.05). Parametric tests (Paired t-tests) were conducted to examine the effect of using AI-powered hydraulic exercise machines on isometric muscle force output following a 6-week intervention. On average, every movement pattern increased in isometric muscle strength from pre- to post-intervention with the exception of trunk flexion, which decreased slightly, though not statically significant (p=0.686). Following Bonferroni correction, significant increases were found for the isometric muscle strength for knee flexion (p=0.002), knee extension (p=0.001), chest press (p=0.001), and leg press (p<0.001), as shown in Table 3.

**Table 3.**
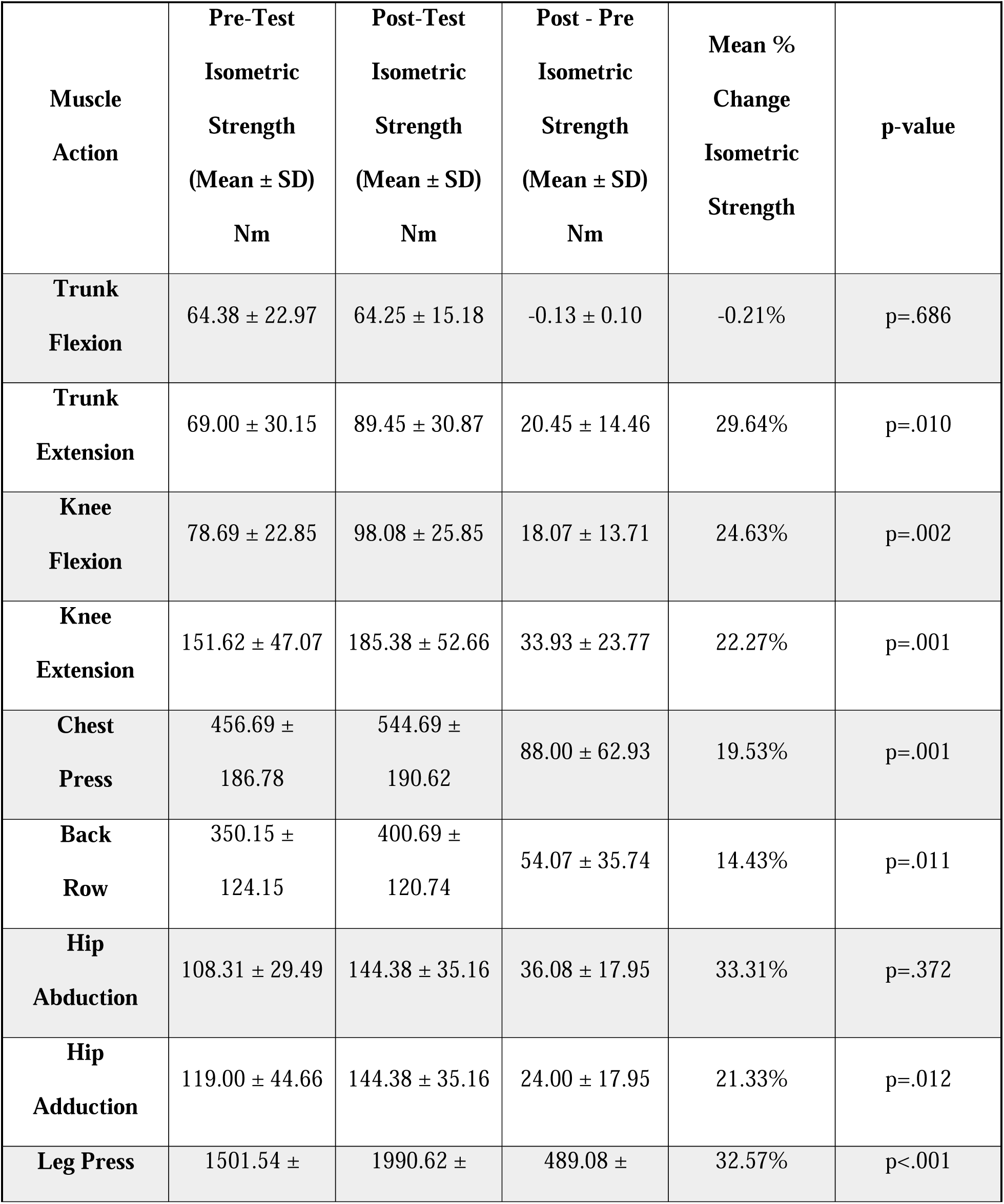
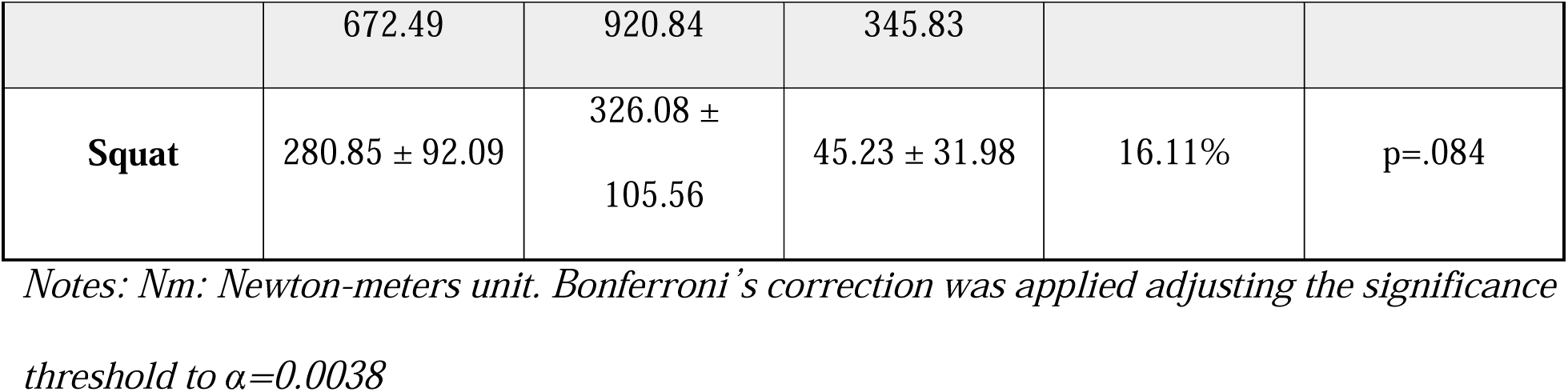
Summary of Pre- and Post-Test Isometric Muscle Strength (Peak Torque) with Statistical Analysis Using Paired T-Test. Table 3 presents a summary of the pre- and post-test means following a 6 week concentric only resistance training program for isometric muscle strength, expressed as peak torque, for each movement, changes in isometric muscle strength (post-minus pre-test values), mean % change from pre- to post-test, and p values following statistical analysis with paired t-test using IBM SPSS Statistics for Windows Version 27. Following the Bonferroni correction, the alpha level was set at 0.0038 for significance. Significant changes in muscle strength were observed in Knee Flexion (p=0.02), Knee Extension (p=0.001), Chest Press (p=0.001) and Leg Press (p<0.001).

### Functional Outcome Measures

The Shapiro Wilk Test indicated that the BBS, TUG, and LEFS data were not normally distributed (p<0.05). Therefore, a non-parametric test (Wilcoxon signed-rank) was used to analyze the data. As illustrated in Table 4, the BBS (p=0.064) and LEFS (p=0.197) average scores increased, while TUG (p=0.901) scores decreased from pre- to post-intervention, however the changes were not statistically significant. The changes were also not clinically significant as they did not meet the MDC or MCID for BBS, LEFS, or TUG.

**Table 4.** Summary of Pre- and Post-Test Outcome Measures with Statistical Analysis Using Wilcoxon Signed Rank Test. Table 4 presents a summary of the pre- and post-test means for functional outcome measures, changes in functional outcome measures of the Berg Balance Scale (BBS), Lower Extremity Functional Scale (LEFS), Timed Up and Go (TUG) (post-minus pre-test values), MDC and MCID for each outcome measure, and p values following a six week intervention period of concentric-only resistance training using hydraulic resistance training equipment. Statistical analysis was performed with the Wilcoxon Signed Rank test using IBM SPSS Statistics for Windows Version 27 was performed. TUG scores are noted in seconds, and a negative value in post-pre reflects an improvement. No statistically significant differences were observed (p>0.0038).

| Outcome | Pre-Test<br>(Mean ± SD) | Post-Test<br>(Mean ± SD) | Post - Pre<br>(Mean ± SD) | MDC | MCID | p-value |
| --- | --- | --- | --- | --- | --- | --- |
| BBS | 52.46 ± 4.75 | 53.77 ± 2.74 | 1.31 ± 0.92 | 6.5<sup>23</sup> | 5<sup>24</sup> | 0.064 |
| LEFS | 67.38 ± 10.14 | 70.46 ± 10.36 | 3.08 ± 2.18 | 9<sup>21</sup> | 9<sup>21</sup> | 0.197 |
| TUG (s) | 10.20 ± 2.93s | 10.18 ± 3.14s | -0.02 ± 0.01s | -3s<sup>26</sup> | -3.4s to -<br>3.5s<sup>27,28</sup> | 0.901 |

*Notes: BBS: Berg Balance Scale, LEFS: Lower Extremity Functional Scale , TUG: Timed-Up and Go, MDC: Minimal Detectable change, MCID: Minimal Clinically Important Difference, s: time in seconds. Bonferroni's correction was applied adjusting the significance threshold to $\alpha=0.0038$*

## DISCUSSION

This study investigated the effects of a 6-week concentric only resistance training protocol using HRT machines on the balance, perceived lower extremity function, fall risk, and muscle strength in older adults. Significant improvements were observed in isometric muscle strength for knee flexion, knee extension, chest press, and leg press. Although positive trends were seen on the BBS, LEFS, and TUG, these changes did not reach statistical or clinical significance. Age-related declines in muscle strength and power have been widely documented and can be related to fall risk in the older adult population.^30^ Therefore, the improvements in motor performance observed in this study may have meaningful implications for functional independence and fall prevention, despite the lack of statistical significance on the functional outcome measures.

This study focused on concentric-only resistance training using a protocol of three sets of ten repetitions, primarily targeting hypertrophy of a muscle, rather than endurance. Type II fast twitch muscle fibers, which contribute to power and strength-oriented activities, have been shown to respond more significantly to high intensity resistance training compared to Type I slow twitch muscle fibers.^31^ As humans age, Type II fibers atrophy more than Type I fibers^30^, yet they are crucial for preventing falls in the older population due to their role in rapid force production.^32^ The muscle actions that demonstrated statistical significance, such as leg press and chest press, are commonly found to be used for power and strength activities, likely driven by Type II muscle activity. In contrast, the functional activities that were not statistically significant, such as balance, may be more endurance-oriented or influenced by multiple systems beyond muscle strength.

Aging also leads to shifts in muscle fiber composition, with a preferential loss of Type II fast-twitch fibers, resulting in decreased power output and slower movement speed.^33,34^ Additionally, the aging process leads to motor unit reorganization, characterized by the loss of larger, fast-twitch motor units and compensatory reinnervation by smaller, slow-twitch motor units, which alters the efficiency and speed of muscle contractions.^35^ Furthermore, older adults exhibit lower motor unit discharge rates during maximal voluntary contractions, reducing the rapid activation of muscles and impairing overall muscle performance.^36^ Decreased fast-twitch fiber recruitment leads to reduced power and speed,^35^ impairing the effectiveness of the steppage and hip strategies needed to rapidly counteract balance disturbances and prevent falls.^37^

Traditional whole body resistance training has been shown to increase muscle mass, strength and performance in older adults.^38^ Both 6-week and 12-week studies have shown increased benefits for older adults participating in resistance training, supporting improvements in daily functional capacity.^38^ Resistance training has also been shown to enhance gait parameters, such as straight-line walking speed, step length, and overall gait performance.^9^ Kudiarasu et al. compared the effects of concentric and eccentric resistance training on adults with Type II Diabetes. With regard to the TUG, only the eccentric training group improved significantly.^39^ Other studies have shown significant improvements on TUG with concentric training, however, the intervention involved was treadmill training rather than utilizing exercise machines, which is more closely related to the activities involved in the TUG.^12^ Being that our study focused on concentric resistance training only, this could be a contributing factor as to why significant improvements were not seen on the TUG.

While traditional resistance training in the older adult population has been widely studied, minimal research has been conducted on the effect of HRT. Lee et al.^15^ conducted a 12- week study utilizing concentric only resistance training using HRT machines and, similar to the results of the present study, observed significant changes in peak torque for knee flexion, knee extension and chest press. In contrast to the present study, Lee et al.^15^ also found significant changes in peak torque for trunk flexion, trunk extension, and chest pull/row. The lack of significance for these movements in the present study could be due to the shorter training period.

Increases in knee flexion and extension motor performance may have implications for balance and function in older adults. Balance and mobility impairments, decreased muscle strength and decreased agility have been shown to be predictors of falls in older adults.^40,41^ Aging leads to declines in sensory structures, including the preferential loss of distal large myelinated sensory fibers and receptors, and impaired lower-extremity proprioception, vibration sense, discriminative touch, and balance.^42^ Proprioceptive input plays a crucial role in maintaining balance by providing continuous feedback on body position and movement.^43^ Therefore, along with the loss of fast-twitch muscle fibers and decreased neuromuscular coordination, these sensory deficits result in delayed reaction times, impairing the ability to recover from perturbations.

It has been shown that older adults utilize lower body co-contraction and joint stiffness to improve stability in standing and co-contraction is greater in those with less balance stability.^44,45^ Chandran et al.^46^ found that older adults have higher co-contraction of knee muscles during stair climbing and gait compared to younger adults. Marshall et al.^47^ evaluated quadriceps EMG activity during stair climbing and ambulation. They found that while older adults had significantly lower peak quadriceps muscle EMG activity during maximum voluntary contraction compared to younger adults, they exhibited significantly higher EMG activity during these functional activities. It was also noted that the biceps femoris muscle was co-activating as an antagonist muscle to assist in maintaining joint stability to reduce fall risk.^47^ Given that knee flexion and knee extension strength are important for functional tasks such as stair climbing and ambulation and co-contraction of these muscles is used as a balance strategy, the significant improvements observed in these muscle groups suggest that concentric only resistance training using HRT machines can be a promising intervention for maintaining functional independence as we age.

Knerl et al.^48^ conducted a 6-week study on the effects of strength training, balance training, and combined strength and balance training on dynamic balance in older adults, using a training frequency of twice per week. Similar to our findings, positive trends were noted on balance tests without statistical significance. This suggests an intervention frequency of twice per week over 6 weeks may be insufficient to elicit significant changes on balance tests in older adults. More research is needed to determine the optimal HRT protocol for improving balance and decreasing fall risk in this population.

The individuals in this study were healthy, independent, community-dwelling older adults with baseline scores that indicated a relatively low risk of falls.^21,22,25^ Available MDCs and MCID values for BBS and TUG are based on populations that are at a higher risk of falling or patients post-stroke. Reported MDCs include 6.5 points on BBS,^23^ 9 points on LEFS,^21^ and 3 second improvement on TUG^26^. Available MCIDs include 5 points on BBS,^24^ 9 points on LEFS^21^, and 3.4-3.5 second improvement on TUG^28^. Participant baseline scores on BBS (mean: 52.46) and on TUG (mean: 10.20s) suggest a potential ceiling effect. While positive trends were noted for all outcome measures, this likely contributed to the inability to detect statistically significant improvements or reach thresholds for MDC and MCID.

### Limitations

This study had some limitations. First, attendance was not strictly controlled. Only three participants attended the full 12 sessions over 6 weeks and 9 attended fewer than 12. As members of the senior center, participants had unrestricted access to the machines and could train more frequently than the prescribed 2 times per week. These factors introduced variability in training exposure. Second, participation in outside activities was not monitored. These external activities could have potentially confounded the results making it difficult to isolate the effects of the HRT protocol. Third, the study recruited healthy individuals who were already at a relatively low risk of falling. This may have contributed to a ceiling effect limiting the ability to detect significant changes or to meet the MDC and MCID thresholds for the BBS, LEFS, and TUG.

To address these limitations, future studies should include stricter protocols on training frequency, limit participation in outside activities, and consider including participants with lower baseline scores on BBS, TUG, and LEFS to better evaluate the effects of concentric only resistance training protocol using HRT machines on balance, fall risk, and perceived lower extremity function.

## CONCLUSION

In conclusion, the findings of this study suggest that a 6-week concentric only resistance training protocol using HRT machines can effectively improve isometric muscle strength in healthy older adults. Although improvements in balance, fall risk, and perceived lower extremity function did not reach statistical significance, positive trends were noted. Future studies should evaluate the effectiveness of a concentric only resistance training protocol on individuals with lower baseline scores on BBS, TUG, and LEFS. Overall, concentric resistance training using HRT machines may be a valuable intervention for improving motor performance and maintaining functional independence in an aging population.

## ACKNOWLEDGMENTS

We would like to thank Dr. Maria Knikou, Dr. Zaghloul Ahmed, and Dr. Wei Zhang for their guidance on this project. We appreciate your support.

## DATA AVAILABILITY STATEMENT

The data that support the findings of this study are available from the corresponding author, AR, upon reasonable request.

